# Tracking cross-border transmission of Rwanda’s successful dominant rifampicin-resistant Mycobacterium tuberculosis clone using genomic markers

**DOI:** 10.64898/2026.03.29.26349652

**Authors:** Isabel Cuella-Martin, Wim Mulders, Jelle Keysers, Francois Hakizayezu, Hosee Niyompano, Docteur Runyambo, Willem Bram De Rijk, Jody Phelan, Yves Mucyo Habimana, Patrick Migambi, Michel Sawadogo, Claude Mambo Muvunyi, Bouke C. de Jong, Jean Claude Semuto Ngabonziza, Leen Rigouts, Conor J. Meehan

## Abstract

**Background:** In Rwanda, genomic surveillance identified a dominant multidrug-resistant tuberculosis (MDR-TB) strain, the R3clone, responsible for approximately 70% of rifampicin-resistant TB cases. Its presence beyond Rwanda remains unexplored.

**Methods:** Unique genetic signatures of the R3clone were defined using whole-genome sequencing of MDR-TB isolates from Rwanda. We developed a targeted qPCR assay detecting a clone-specific single-nucleotide polymorphism. With these tools, we screened isolates from neighbouring countries and public genomic repositories.

**Results:** We identified 375 R3clone isolates, including 264 from historical Rwandan collections (1991-2021), 49 from recent Rwandan diagnostic routine (2021-2024), 25 from historical Burundi isolates (2002-2013), and 37 among public repositories from several countries. The R3clone-specific qPCR showed 100% specificity in distinguishing the R3clone from other MTBC (sub-)lineages. Transmission analysis revealed cross-border transmission of the R3clone within the Great Lakes Region.

**Conclusion:** This study comprehensively assesses cross-border transmission of a dominant MDR-TB strain, highlighting the need for coordinated international surveillance.

**Impact statement:** Tuberculosis management is primarily undertaken at a national level, with associated contact tracing and public health interventions also being performed within country. Several countries have signature strain or drug resistance patterns within their *Mycobacterium tuberculosis* population, allowing for tracking of circulating clones within their borders. Rwanda is one such case, with the R3clone dominating the MDR-TB cases within the country.

Previous studies of the R3clone have shown that improved patient management and treatment can reduce the prevalence of this clone within Rwanda. Our study confirms this decline in population size of the R3clone within Rwanda. We also show that this is not a Rwanda-restricted clone; by developing a low-cost qPCR-based detection method we uncovered this clone in Burundi and through extended search of online databases additionally found it in other neighbouring countries.

Our findings demonstrate that tracking of *M. tuberculosis* needs to extend beyond country borders, especially in endemic regions with high levels of inter-country migration. Cross-border tracking and co-operation will enhance intervention measures and reduce cases of MDR-TB transmission.

**Data summary:** All samples sequenced in this project from the InnoR3Tb project (Rwanda and Burundi) are available via the ENA projects PRJEB106304 and PRJEB95959. The exact ENA accession codes for each sample are listed in Table S3 alongside sampling dates and countries.

## Background

Tuberculosis (TB) remains a significant global public health issue, with the emergence and spread of multidrug-resistant TB (MDR-TB), resistant to both rifampicin (RIF) and isoniazid (INH), undermining TB control progress.^1^ Understanding MDR-TB transmission dynamics, like the circulation and dissemination of predominant clones, is essential for control strategies. Cross-border transmission of MDR-TB has been documented between neighbouring countries and even across continents along migration routes.^2–5^ However, current MDR-TB control efforts remain largely constrained within national boundaries.

In Rwanda, genomic surveillance identified a dominant clone of the Ugandan sub-lineage 4.6.1.2, the Rwanda rifampicin-resistant clone (R3clone).^6^ This clone, responsible for approximately 70% of rifampicin-resistant TB (RR-TB) in Rwanda, appears to have driven the RR-TB epidemic from the 1990s through the 2000s. Although its prevalence seems to decrease since 2014, the R3clone remains significant due to its full resistance to first-line TB drugs and the carriage of Ser450Leu mutation in the *rpoB* gene, a mutation associated with rifampicin resistance and minimal fitness cost.^7^ While well-characterised within Rwanda, the R3clone presence beyond national borders remains largely unexplored.

Rwanda sits within the Great Lakes region of East and Central Africa and primarily consists of Burundi, The Democratic Republic of Congo (DRC), Rwanda, Tanzania and Uganda. In the Great Lakes Region, where population movement is common, cross-border transmission might pose a challenge to MDR-TB control, especially with uneven surveillance systems and limited genomic data.^8,9^

Traditional molecular epidemiological methods, such as spoligotyping, have been used to assign isolates to shared international types (SITs) and lineage families.^10^ Although useful for molecular epidemiology at broad scales, spoligotyping lacks the discriminatory power to define individual transmission chains.^11^ Whole genome sequencing (WGS) has become the gold standard for high-resolution MDR-TB genomic analysis. However, its cost (especially compared to spoligotyping), infrastructural requirements, and limited availability in many high-burden countries restrict its widespread use in routine surveillance.^12,13^ Tracking specific successful clones like the R3clone is epidemiologically important as their population dynamics serve as markers of drug-resistant TB control effectiveness and can guide targeted interventions. Affordable and scalable molecular tools that can detect the R3clone among other circulating strains would significantly improve targeted enhanced case finding efforts.

In this study, we defined a unique genetic signature for the R3clone identification through targeted molecular diagnostics. Using this definition, we screened isolates from neighbouring countries and public genomic databases to explore the geographic extent and cross-border transmission potential of the clone.

## Methods

### Rwanda dataset

The Rwanda dataset included retrospective historical isolates and prospective recently collected samples. Retrospective data consisted of RR-TB isolates collected nationwide for diagnostic purposes between 1991 and May 2021.^14^ Prospective nationwide isolates from the diagnostic cohort study “InnoR3TB” included all consecutive RR-TB patients from May 2021 through April 2024.

Sample collection, processing, culture and genomic extraction were performed as previously detailed.^6,15,16^ WGS was outsourced to FISABIO (Spain), KU Leuven (Belgium), CD Genomics (USA) and MicrobesNG (UK), on an Illumina HiSeq or NovaSeq 6000 platform.

### Reconstruction of the R3clone phylodynamics within Rwanda

Non-MTBC reads were excluded from the raw Fastq dataset using Centrifuge v1.0.4, retaining only reads aligning to MTBC variant.^17^ High-quality reads were mapped to the inferred ancestral *M. tuberculosis* genome using MTBseq v.1.1.0 with default parameters, with a 5% variant frequency cut-off for reliable variants.^18,19^

A single-nucleotide polymorphism (SNP) alignment was created via MTBseq, and constant site pattern counts were estimated using custom Python scripts available at https://github.com/conmeehan/pathophy. To define R3clone membership, we applied a loose 12-SNP cut-off, using historical Rwandan R3clone genomes as a reference. Population size estimates were undertaken using BEAST v1.10.4 from this SNP alignment with constant site count correction.^20^ BEAUti was used to create the XML file for BEAST with the following parameters: tip dates were added based on the date of isolation; where only the isolation year was known, the uncertainty was set to 1 to estimate over the entire year. A GTR+G site model was used, along with a lognormal, uncorrelated, relaxed clock and a time-aware GMRF Skyride tree prior.^21,22^ The ucld.mean was set to 1/x with a starting value of 3.5^-9^ to match the settings and estimates of previous analyses.^23^

The Markov chain Monte Carlo (MCMC) was set to a length of 1B, sampling every 100,000 steps with four independent runs to assess convergence. Both logs and trees from were combined using LogCombiner with a 50% burn-in per run. Tracer v.17.1 was used to check convergence and construct the Skyride plot; the youngest age was set to 2024, corresponding to the most recently isolated.^21^

### Creating an R3clone-specific identifying genomic profile

To enable R3clone identification without WGS, we used three complementary approaches:

#### 1) Drug resistance mutation pattern

Previously reported resistance-associated mutations in the R3clone, particularly *rpoB* Ser450Leu and *katG* Ser315Thr, were used as reference markers.^6^ All genomes were analysed using TB-Profiler v6.2.1 with default settings.^24^

#### 2) Spoligotype pattern

Spolpred2, implemented within TB-Profiler, determined spoligotype pattern of all retrospective Rwandan strains.^25^ Samples where the spoligotype pattern was all 0’s were removed. After exclusion of samples with insufficient coverage, 23 spoligotype patterns were identified among R3clone isolates, collectively defined as the R3clone spoligotype family (R3-family) (Table S1).

#### 3) Clone-specific SNP

Genomes of the R3clone group and non-R3clone Rwanda group were compared to find an R3clone-specific SNP. The SNP alignment created was converted to variant call format (VCF) file using SNP-sites v2.5.1 and converted to a diploid format using haploid2diploid.py (github.com/jodyphelan/haploid2diploid.py).^26^ The resulting VCF file was input to VCFtools v0.1.16 with the two Rwanda groups also input via the --weir-fst-pop flags.^27^

To confirm the uniqueness of the selected SNP, all MTBC raw Illumina data from the Sequence Read Archive (SRA) was retrieved (February 2025; n=189,326). Genome-VCF files were created using bwa mem, GATK and samtools.^28–30^ The corresponding VCF files from this dataset were queried to look for the clone-specific SNP identified above. A SNP distance matrix was created between all isolates which had the clone-specific SNP using the setProcessPooledVCF.pl function of the LYVE-SET toolkit. A quantitative polymerase chain reaction (qPCR) assay targeting this R3clone-specific SNP was developed and validated (Supplementary material).

### Selection of Burundi samples

Between 1999 and 2013, 192 RR-TB isolates were collected in Burundi through routine surveillance by the Damien Foundation and stored at ITM. After excluding 49 duplicates, the remaining 143 unique isolates were examined. Sanger sequencing of the *rpoB* gene and spoligotyping were performed to screen for genetic profiles and spoligotyping patterns.^10,31^ Isolates identified as possible R3clone were subsequently subjected to the R3clone-specific SNP qPCR as described (Supplementary material). Samples that were positive in the R3clone-qPCR underwent WGS by Illumina HiSeq X Ten (CD Genomics, USA).

### Selection of global public genomic data for transmission analysis

The retrieved MTBC raw Illumina data from the SRA were filtered by lineage. 779 isolates from lineage 4.6.1.2 were explored further. In total, 267 isolates from the historical Rwanda project (PRJEB43270) were removed, as already included in the Rwanda retrospective historical dataset, along with 37 single-end isolates. The country of origin and sample identifiers were evaluated to identify and remove 174 duplicate submissions.

### Whole genome sequencing, clustering and phylogeny

The SNP-calling pipeline used for the Bayesian phylodynamics was applied to all R3clone raw FastQ files, regardless of their dataset origin. Drug-resistance profiles were called using TB-Profiler v6.2.1 with the tbdb mutation database.^24^ Samples were excluded if they represented polyclonal TB infections.

A SNP alignment was generated using MTBseq, and constant sites across the remainder of the genome were calculated using a custom Python script.^6,19^ To determine inclusion in the R3clone cluster, a 12-SNP cut-off was applied as per established criteria. A minimum spanning network was constructed using PopART v1.7, using default parameters with branch lengths representing SNP distances between isolates. The network was annotated by country of origin. Aligned R3clone SNP data were also employed to build a Maximum Likelihood phylogeny with RAxML-NG v1.2.0.^32^ The run used a GTR+G+ASC substitution model with site-repeat optimisation and all invariant sites included as counts per nucleotide (A/C/G/T), incorporating 10 starting trees and 200 bootstrap replicates for stable tree construction.^33^ The phylogenetic tree was annotated using the Interactive Tree Of Life (iTOL v6) software.^34^

### Ethics

The study protocol was approved by the Rwanda National Ethics Committee, Kigali, Rwanda (IRB 00001497 of IORG0001100; Ref No·705/RNEC/2021), the Institutional Review Board of the Institute of Tropical Medicine, Antwerp, Belgium (IRB/AB/AC/157; Ref No.1525/21; 23/09/2021), and the Ethics Committee of the Antwerp University Hospital, Belgium (REG No. B3002021000230; 22/11/2021).

## Results

### Rwanda R3clone population overview and dynamics

Our analysis incorporated both the historical R3clone dataset (1991-2021) and newly enrolled isolates from the InnoR3TB study period (2021-2024) (Figure 1). The historical collection, previously described in detail, comprised 264 R3clone isolates collected nationwide over a three-decade period.^6,14^

**Figure 1.**
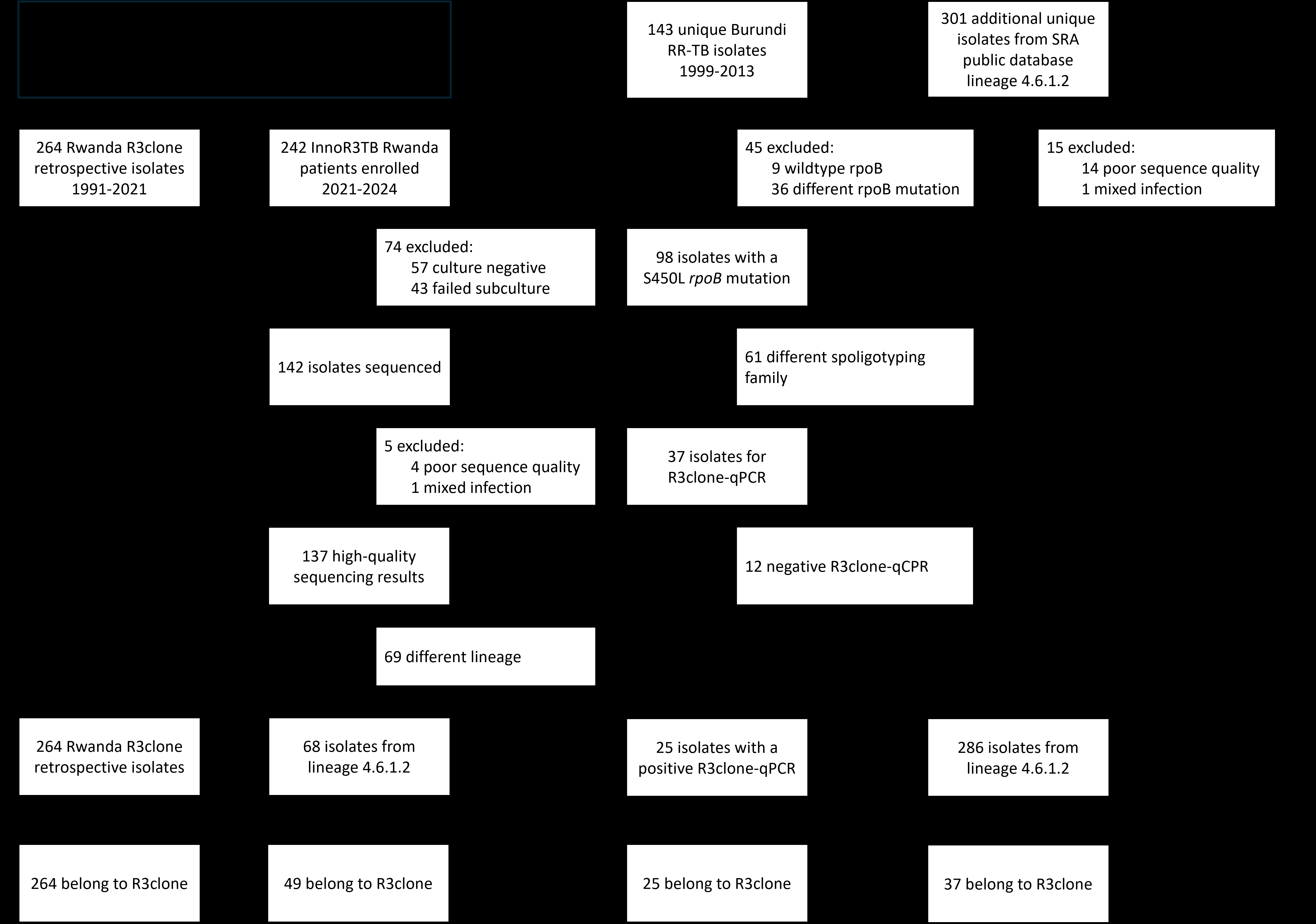
Flowchart of study population, exclusion criteria, and inclusion in transmission clustering analysis. RR-TB: rifampicin-resistant tuberculosis. SRA: Sequence Read Archive. R3clone: Rwanda rifampicin-resistant clone. qPCR: quantitative polymerase chain reaction.

During the InnoR3TB study period in Rwanda, 242 patients were enrolled nationwide from May 2021 to December 2024. Of these, 185 (76.4%) were culture positive, and 142 were successfully subcultured and sent for sequencing. After excluding four isolates with poor sequence quality and one with mixed populations, we analysed 137 patients with high-quality sequencing results. The demographic profile revealed a male predominance (114, 83.2%) with a median age of 37 years (IQR: 28-47 years). HIV co-infection was present in 22 patients (16.1%). Most patients (108, 78.8%) were new TB cases, with only 29 (21.2%) having previous TB treatment history. Geographic distribution showed that approximately one-third of patients came from Kigali province (49, 36.0%), another third from Eastern province (41, 30.1%), with the remainder distributed across Southern (20, 14.7%), Western (16, 11.8%), and Northern (10, 7.4%) provinces of Rwanda.

Lineage analysis revealed a diverse distribution, with Ugandan sub-lineage 4.6.1.2 representing 49.6% (68/137) of isolates. Other lineages included 4.6.1.1 (25, 18.2%), 4.7 (17, 12.4%), 4.3 (14, 10.2%), with smaller numbers of 4.8 (3, 2.2%) 4.1 (2, 1.5%), 4.2 (2, 1.5%), and 4.4 (1, 0.7%), while five (3.6%) were lineage 3 (CAS). The 68 isolates from the lineage 4.6.1.2 were included in the later transmission analysis.

An updated overview of the R3clone population dynamics in Rwanda, using the expanded dataset, confirmed the expansion of the clone in Rwanda from the 1980s through to the early 2000s, followed by a levelling off in subsequent decades (Figure 2). From 2014 onwards, there appears to be a decline in the clone population

**Figure 2.**
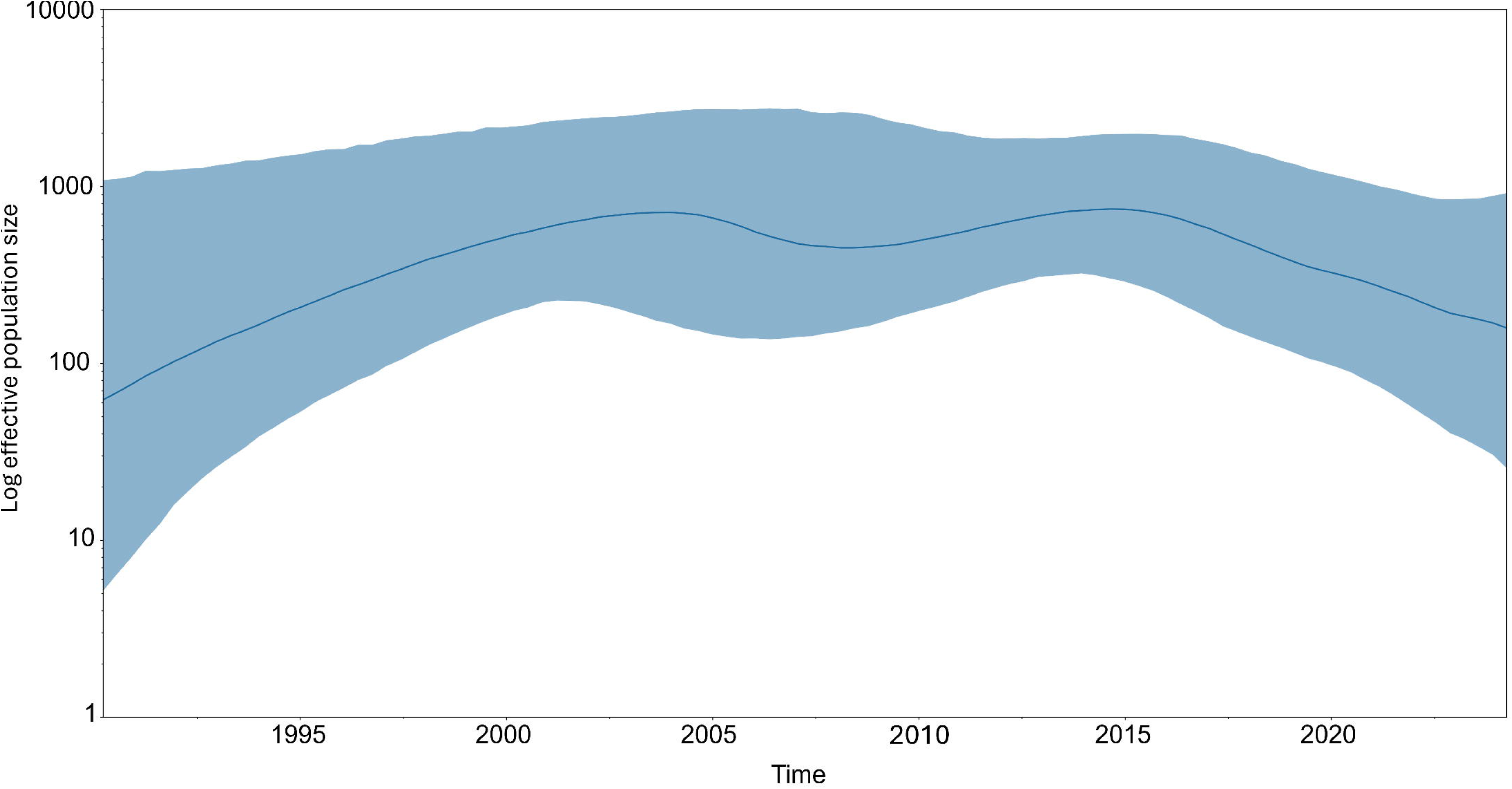
Population dynamics of R3clone in Rwanda until 2024. The solid blue line indicates the mean population size. The shaded blue area indicates the 95% high-posterior-density interval estimate.

### Development of an R3clone-specific qPCR

The population genetics-based approach identified six SNPs with an fixation index (F_ST_) of one between the R3clone and non-R3clone Rwanda isolates (Table S2). The C25631G SNP was selected as the R3clone-specific marker because it is intergenic, between Rv0020c and Rv0021c, whereas all others are within genes (Table S2), potentially subject to greater evolutionary pressure. This SNP was searched in the SRA MTBC dataset and was not present in any strain that was not part of the R3Clone cluster.

### Genotypic characterisation and selection of Burundi isolates

Parallel spoligotyping and Sanger sequencing of the *rpoB* gene were performed on the 143 unique RR-TB isolates from Burundi archived at ITM (Figure 1). No single spoligotype pattern was observed but almost all had the distinguishing 1111111100001110111 in spaces 37-43 (Table S1), marking them as belonging to SIT53. Sanger sequencing revealed that 68.5% (98/143) carried the Ser450Leu mutation, while 6.3% (9/143) had wild-type *rpoB* sequences and 25.2% (36/143) harboured other resistance-associated *rpoB* mutations.

Spoligotyping identified eight distinct families, with 29 isolates remaining unclassified. The Latin American-Mediterranean (LAM) family predominated (n=71, 49.7%), followed by the R3-family (n=18, 12.6%) (Table 1). Within the R3-family isolates, 15 carried the Ser450Leu mutation, one had Leu452Pro, and two exhibited wild-type *rpoB* sequences.

**Table 1.**
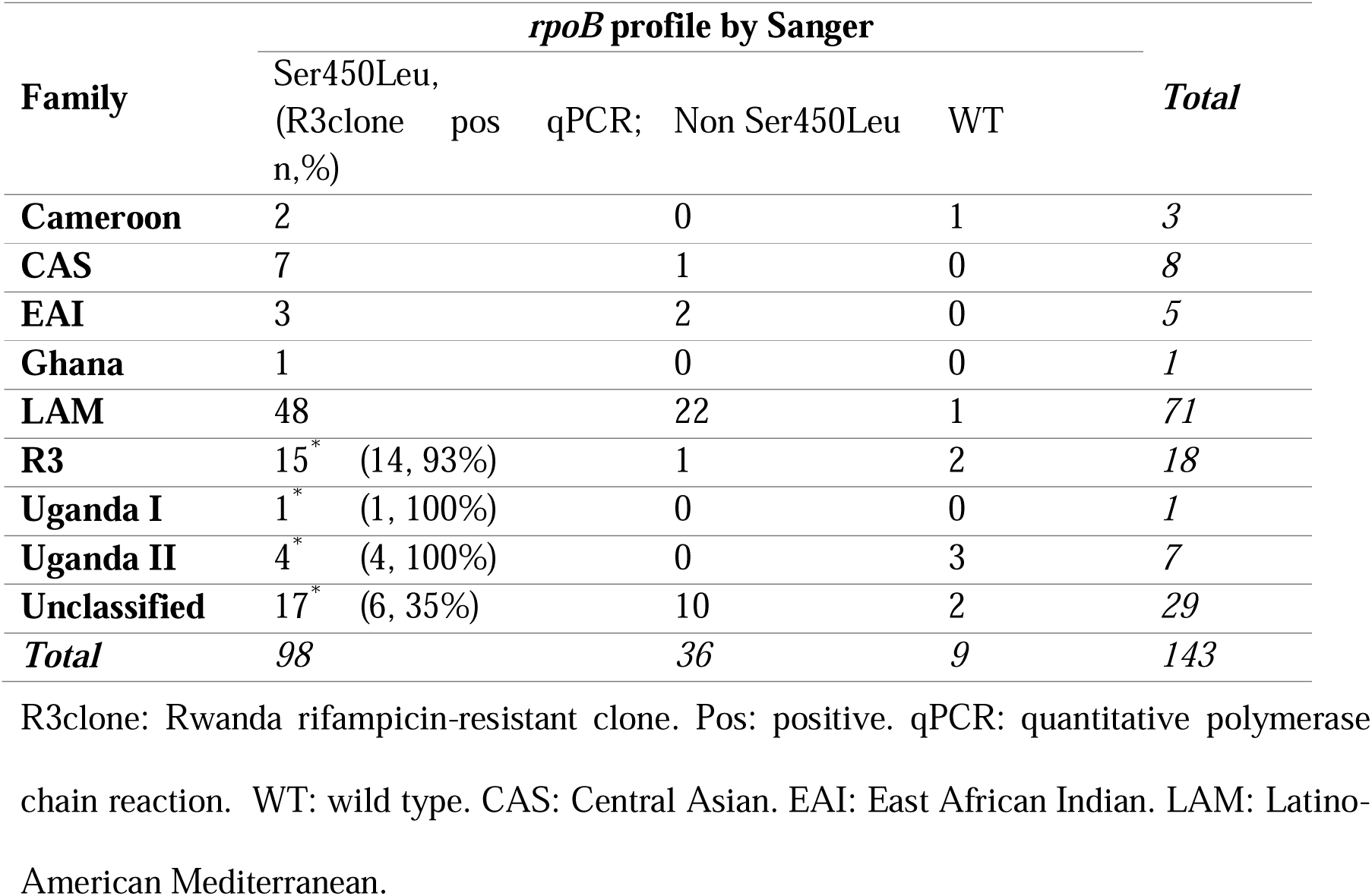
Distribution of spoligotype families and corresponding rpoB mutation profiles among 143 RR-TB isolates from Burundi. *Isolates on which R3clone-qPCR was performed on.

Based on the presence of the Ser450Leu mutation and genetic relatedness, a subset of 37 isolates was selected for R3clone-qPCR testing. This subset comprised all 15 R3-family isolates with Ser450Leu, as well as additional isolates from Uganda I (n=1), Uganda II (n=4), and unclassified families (n=17) that also harboured Ser450Leu.

All 37 isolates that underwent R3clone-qPCR were amplified with the TBc probe, confirming the presence of MTBC DNA. Among them, 25 (67.6%) tested positive for the R3clone-specific SNP, with cycle threshold (Ct) values ranging from 21 to 27. These isolates were distributed across the R3-family (n=14), Uganda I (n=1), Uganda II (n=4), and unclassified lineages (n=6). The 25 R3clone-qPCR positive samples proceed to subsequent transmission analysis; WGS was not performed on the other samples.

### Transmission clustering analysis

A total of 643 isolates, either identified as historic R3clone, belonging to L4.6.1.2, or as positive at R3-qPCR, were included in the transmission clustering analysis (Figure 1). Using a 12-SNP threshold and a loose clustering definition, 624 isolates (97.0%) grouped into 27 genomic clusters. The largest cluster contained 394 isolates, which included the historic R3clone. The second largest cluster consisted of 54 isolates, while the remaining 25 clusters ranged from 2 to 43 isolates.

Among the 394 isolates initially grouped within the R3clone cluster, 18 isolates from the public SRA database showed *rpoB* mutation profiles inconsistent with the R3clone-defining pattern: 11 were wild-type *rpoB*, two carried the Asp435Val, and five had mutations at codon 445 (two His445Asp, one His445Leu and two His445Tyr). One isolate also showed a discordant *katG* mutation, carrying an Asp735Ala mutation of uncertain significance instead of the characteristic Ser315Thr. Phylogenetic analysis revealed that these 19 isolates were basal to the central cluster, suggesting that *rpoB* and *katG* mutations are essential for R3clone definition and that a SNP cut-off alone might not be sufficient. After excluding these 19 basal isolates, 375 were defined as actual members of the R3clone (Figure 3). These comprised 264 (70.4%) isolates from the retrospective Rwanda collection, 49 (13.0%) from the InnoR3TB cohort, 25 (6.7%) from the Burundi study, and 37 (9.9%) from the SRA database (Figure 1).

**Figure 3.**
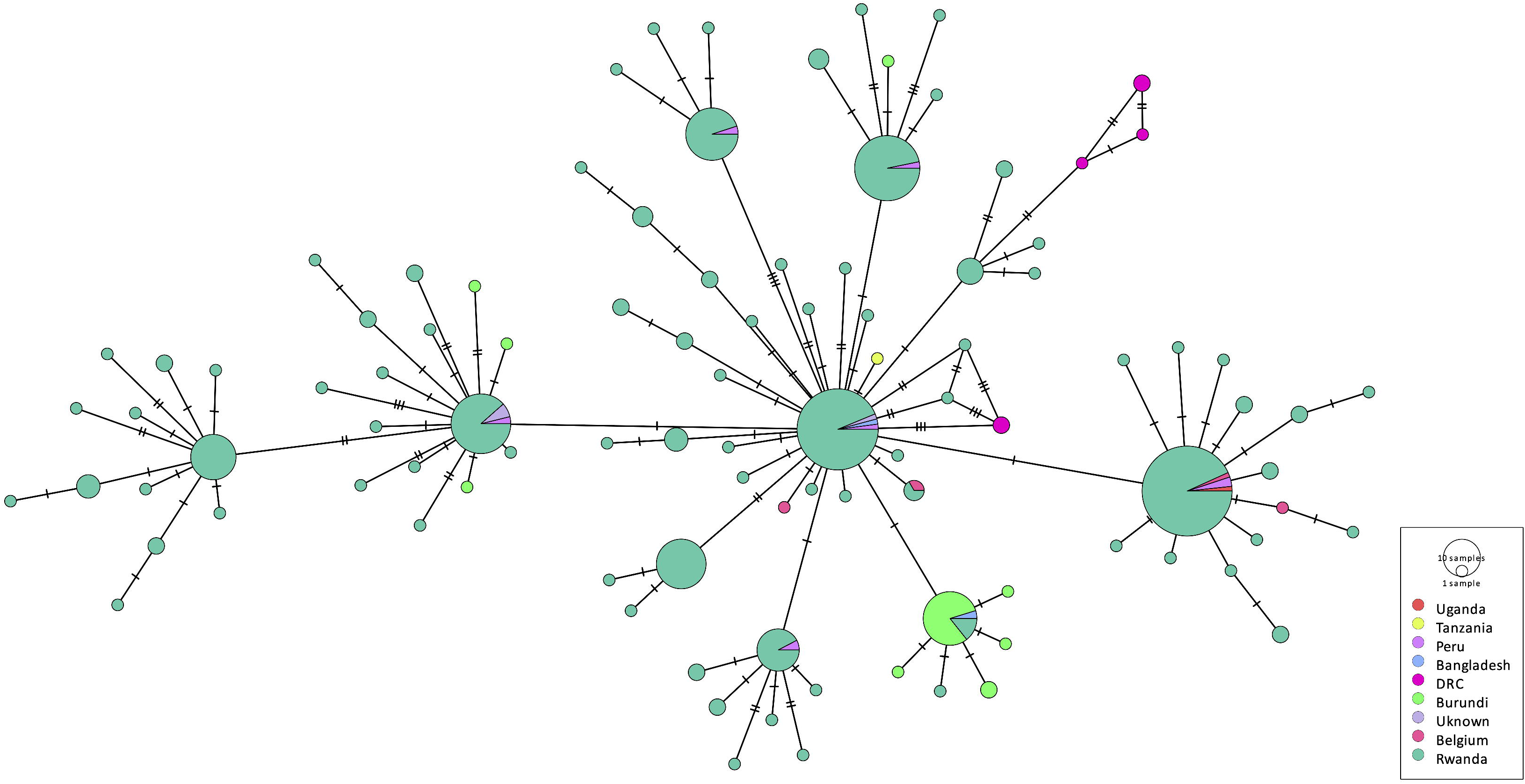
Minimum spanning tree of 375 confirmed R3clone isolates. Nodes are colored by country of origin and sized proportionally to the number of isolates with identical genotypes. Branch lengths represent SNP distances.

SRA-derived R3clone isolates originated from countries bordering Rwanda, including the Democratic Republic of Congo (DRC, n=6), Burundi (n=1), Uganda (n=1), and Tanzania (n=1), as well as from more distant locations such as Bangladesh, Belgium, and Peru (Table 2).

**Table 2.** Isolates from the SRA database with a Ser450Leu rpoB mutation that were included in the transmission analysis, stratified by either inclusion or not in the R3clone cluster and country of origin.

| Country of origin | 5 SNPs cut-off |  |  | *Total* |
| --- | --- | --- | --- | --- |
|  | R3clone |  | Non-R3clone |  |
|  | n,(%) | Approximate date of isolation | n,(%) |  |
| **Bangladesh** | 2 (100%) | 1995, 2000 | 0 | 2 |
| **Belgium** | 4 (100%) | 2007-2019 | 0 | 4 |
| **Burundi** | 1 (100%) | 2002 | 0 | 1 |
| **Democratic Republic of Congo** | 6 (17%) | 2005-2010 | 30 (83%) | 36 |
| **Kenya** | 0 | - | 1 (100%) | 1 |
| **Netherlands** | 0 | - | 1 (100%) | 1 |
| **Peru** | 7 (88%) | 2012 | 1 (12%) | 8 |
| **Rwanda** | 12 (100%) | 2003-2004 | 0 | 12 |
| **Tanzania** | 1 (100%) | 2014 | 0 | 1 |
| **Tunisia** | 0 | - | 1 (100%) | 1 |
| **Uganda** | 1 (6%) | 2003-2007 | 15 (94%) | 16 |
| **United Kingdom** | 0 | - | 1 (100%) | 1 |
| **Unknown** | 3 (60%) | - | 2 (40) | 5 |
| **Total** | 37 (42%) | - | 52 (58%) | 89 |

SNPs: single-nucleotide polymorphisms. R3clone: Rwanda rifampicin-resistant clone.

All 375 confirmed R3clone isolates harboured the characteristics of the katG Ser315Thr mutation associated with isoniazid resistance and the rpoC Pro481Thr compensatory mutation. Regarding ethambutol resistance, 369 isolates (98.4%) carried either the embB Met306Val or His1002Arg mutation, both of which have been found previously to be associated with the R3clone. The remaining six isolates (2.6%), all from DRC, carried other embB profiles: three were wild-type, two had Asp1024Asn, and one had Asp354Ala mutations. These six isolates from DRC had wild-type pncA sequences. Only four additional isolates from early Rwanda samples (1991–1993) had also wild-type pncA. (Figure S1).

Among the 26 total R3clone isolates from Burundi – 25 from R3clone-qPCR and one from the SRA database-two distinct subgroups were identified. The larger subgroup (n=22) shared a characteristic 386_388delATG mutation in the *pncA* gene and an *rpoB* Pro45Leu mutation, not associated with resistance. This same mutation pattern was also observed in one isolate from Bangladesh and six from Rwanda collected between 2015 and 2018. The remaining four Burundian isolates carried either a *pncA* Gln10Arg (n=1) or His43Pro (n=3) mutation and lacked the *rpoB* Pro45Leu.

## Discussion

This study presents the first comprehensive assessment of the geographic distribution of R3clone. By defining unique genetic signatures and screening isolates from neighbouring countries and global databases, we provided evidence of likely cross-border transmission of this MDR-TB strain within the African Great Lakes Region and beyond.

Our previous work showed that R3clone likely originated in the mid-1980s and expanded through the 1990s and 2000s before becoming stationary in the 2010s.^6^ Repeating this phylogenetic analysis with an extended dataset (1991-2024) revealed a continuing decline in population size, extending the downward trend previously observed from 2014.^6^ The sustained reduction aligns with Rwanda’s progressive TB control strategy.^35^ What began as targeted testing for retreatment cases evolved into comprehensive, universal drug susceptibility testing for all bacteriologically confirmed TB patients by 2014, with current data showing that 96% of new TB cases and 94% of previously treated cases underwent rifampicin-resistance testing.^1^ The nationwide implementation of GeneXpert technology has enabled the rapid diagnosis and timely initiation of appropriate treatment, thereby preventing the ongoing transmission of drug-resistant strains.^36^ Additionally, Rwanda’s policy of mandatory hospitalisation for MDR-TB treatment and high treatment success rate for MDR-TB, which exceeded 90% in recent cohorts, have potentially reduced the effective spread of the R3clone compared to previous decades.^1^

Our analysis successfully identified distinctive genomic markers that reliably identify the R3clone, including a signature drug resistance mutation profile and a clone-specific SNP. The presence of the *katG* Ser315Thr and *rpoB* Ser450Leu mutations, along with *rpoC* Pro481Thr, across all R3clone isolates suggests these were foundational mutations acquired early in its evolution.^37^ We demonstrated that the C25631G SNP, located in an intergenic region, is specific to the R3clone and allowed the diagnostic development of the R3clone-qPCR, which represents a significant achievement in molecular diagnostics for MDR-TB surveillance. Our study found the qPCR be 100% specific when tested on a selected cohort from Burundi, where all 25 qPCR-positive isolates were subsequently confirmed as R3clone by transmission analysis. This approach parallels successful SNP-based qPCR technologies developed for other geographically significant MDR-TB strains, such as the Russian Beijing 1071-32-cluster and the Equatorial Guinea MDR strain (EG-MDR).^38,39^

Importantly, our findings indicate that spoligotyping alone is insufficient for accurate R3clone identification. Although spoligotyping initially classified 18 isolates from Burundi (12.6%) as belonging to the R3clone spoligotyping family, subsequent R3clone-qPCR testing revealed that 25 isolates harboured the clone-specific SNP, including some previously classified as Uganda I, Uganda II, and unclassified lineages. This discrepancy highlights the limited discriminatory power of spoligotyping and underscores the need for more comprehensive genomic approaches to accurately track the spread of clonal outbreaks.^11,12^

The identification of R3clone isolates in all four neighbouring countries, particularly Burundi and the DRC, provides evidence of cross-border transmission within the African Great Lakes Region. Our public database exploration suggests R3clone’s presence in additional locations, including Bangladesh, Belgium, and Peru. While SRA submissions may reflect sequencing locations rather than strain origins, international spread through human migration patterns has been reported for other MDR-TB strains, including between high-burden countries and from high-burden to low-burden countries.^40^

Our phylogenetic analysis revealed important evolutionary insights into R3clone. The identification of six DRC isolates with wild-type *pncA* sequences, phylogenetically related to early Rwandan samples (1991-1993), implies that R3clone may have been present in DRC before acquiring additional resistance to pyrazinamide and ethambutol. These findings align with the broader understanding of how MDR-TB clones acquire additional resistance mutations over time, often in response to selective pressure from treatment regimens.^41^ However, the small sample size makes it difficult to confirm in this instance.

In Burundi, we identified two distinct R3clone subgroups, possibly indicating repeated introduction events rather than a single cross-border transmission. The larger subgroup, characterised by the 386_388delATG mutation in *pncA* and by the non-resistance-associated *rpoB* Pro45Leu mutation, was phylogenetically close to Rwandan isolates collected between 2015 and 2018. This suggests ongoing bidirectional movement of MDR-TB strains across the Rwanda-Burundi border, consistent with substantial population movement between these two countries.^42^

The distribution pattern of the R3clone across the region shows interesting variation. While prevalent in Rwanda and well-represented in Burundi, the clone appears less dominant in the DRC and Uganda, despite a confirmed presence. This heterogeneous distribution likely reflects sampling bias, as isolates from the DRC and Uganda may predominantly originate from regions distant from Rwanda, such as Kinshasa for the DRC, rather than border areas where cross-border transmission would be most likely. Differences in surveillance coverage, with some countries having more comprehensive national versus regional sampling, may contribute to this observed variation.^43^

To facilitate practical implementation in resource-limited settings, we propose leveraging the existing diagnostic platforms for R3clone surveillance. The characteristic Ser450Leu *rpoB* mutation produces a distinctive melting temperature pattern in the probes rpoB3 and rpoB4 of the Xpert MTB/RIF Ultra assay.^44,45^ This easily identifiable pattern could serve as a cost-effective initial screening tool for potential R3clone cases in settings where Xpert technology is already deployed. Similarly, conventional first-line probe assays (LPAs), such as MTBDRplus, detect both *rpoB* Ser450Leu (*rpoB* MUT3 probe) and *katG* Ser315Thr (*katG* MUT1/MUT2 probes), allowing the identification of potential R3clone candidates.^46^ These candidates could then be validated with the specific qPCR assay, creating a tiered and economically feasible surveillance strategy. Beyond R3clone detection, this approach could support coordinated regional contact tracing.

In an increasingly interconnected world, where the movement of people facilitates the spread of resistant strains, confining MDR-TB control strategies within national borders is an insufficient approach to address this transnational public health threat.^47,48^ The R3clone serves as an essential case study, illustrating a phenomenon likely occurring with other drug-resistant TB strains globally. MDR-TB control programs can be challenged when patients who carry strains with complex resistance profiles cross regional boundaries, as documented in Cameroon.^49^

Our study had some limitations. First, our investigation relied partly on retrospective samples collected through convenience sampling in Burundi, which may have introduced selection bias. Second, we relied on culturable isolates for WGS analysis, which creates additional selection bias as only successfully cultured samples could be sequenced. Third, samples from different countries covered irregular time intervals with limited numbers of isolates and differences in the comprehensiveness of sample collection within big countries. These constraints limited making inferences about transmission directionality.

In conclusion, this study demonstrates the potential of affordable and scalable molecular tools for enhancing the surveillance of a dominant MDR-TB clone in resource-limited settings. This framework could be applied to track other regionally significant drug-resistant TB strains. The identification of cross-border transmission of the R3clone highlights the urgent need for coordinated regional and international efforts in surveillance, case detection, and treatment to effectively combat the spread of successful drug-resistant TB clones.

## Data Availability

All samples sequenced in this project from the InnoR3Tb project (Rwanda and Burundi) are available via the ENA projects PRJEB106304 and PRJEB95959. The exact ENA accession codes for each sample are listed in Table S3 alongside sampling dates and countries.

## Author contribution

ICM, BCdJ, LR, JCSN, and CJM conceived and designed the study. ICM, FH, HN, DR, JK, WBdR, WM, YMH, PM, MS, and CMM contributed to investigation, data collection, and resources. ICM, WM, and JK performed data curation, analysis, and visualization with methodological input from JP and supervision from CJM and JCSN. ICM and CJM wrote the first draft of the manuscript. BCdJ, LR, and JCSN provided overall supervision, funding acquisition, and critical revision of the manuscript. All authors reviewed and approved the final version.

## Acknowledgements

We thank the reviewers at the Rapid Review/Infectious Diseases website for additional comments on the preprint.

