## Supplementary material for "Tracking cross-border transmission of Rwanda’s successful dominant rifampicin-resistant Mycobacterium tuberculosis clone using genomic markers"

**R3clone-qPCR development and specificity analysis**

The qPCR used custom primers (F: 5’-catggtatcgggacagtgtgagcgag-3’; R: 5’-ggcgtgcgctagccgttcc-3’) and two fluorescent probes: an R3clone-specific probe (5’-aggtatggtcGtccggttgt-3’, FAM-labelled with EDQ quencher) and a TB-complex-specific probe (5’-cgacgacgatgtcggtcacgactcgtc-3’, DFO-labelled with BHQ2 quencher). Each 15 µL reaction included 10 µL of qPCR mastermix (SensiMix™ II Probe mix or GoTaq® Probe qPCR Master Mix), 0.4 µL of each probe (5 µM), 0.44 µL of each primer (18 µM), 0.24 µL of ROX/CXR, 3.48 µL of nuclease-free water, and 5 µL of genomic DNA. Reactions were run on StepOnePlus or QuantStudio 5 systems with the following cycling protocol: initial denaturation at 95°C for 10 minutes, followed by 50 cycles of 95°C for 20 seconds and 66°C for 1 minute. Each run included a positive control (R3clone DNA), a negative control (non-R3clone MTBC DNA), and a no-template control to monitor for amplification specificity and contamination. Samples were considered positive for R3clone if amplification occurred with a threshold cycle (CT) less than 35 and controls behaved as expected.

Two specificity tests were conducted for the R3clone-qPCR assay. The first tested whether the R3 and TBc probes selectively hybridise with R3clone and general MTBC DNA, respectively, without cross-reacting with closely related species. A panel of 121 non-tuberculous mycobacteria (NTM) and 32 closely related non-mycobacterial species was evaluated. The TBc probe yielded positive signals in 7 NTM and one non-mycobacterial isolate. These samples were retested using an IS*6110*-targeted qPCR to determine whether the TBc-probe had reacted with actual MTBC DNA or produced false positives. Six out of eight samples were IS*6110*-positive, suggesting they contained residual MTBC DNA, likely due to employing reusable glass bijoux bottles to prepare bacterial suspensions. The two IS*6110*-negative samples were not investigated further.

The second test evaluated the probe specificity within the MTBC. A clinical reference panel representing MTBC lineages 1 through 10 (L1–L10) was used to verify that the TBc probe detects all lineages and that the R3 probe does not cross-react with non-R3 clone strains. All 25 isolates tested positive with the TBc probe, and none with the R3 probe, confirming lineage-wide specificity.

Finally, the detection limit was assessed using serial dilutions of an R3clone isolate and the H37Rv strain. Results showed that a minimum of 1 whole genome equivalent per microliter was required for consistent detection in all wells by both probes.

Table S1.Spoligotype patterns identified among R3clone isolates, collectively defined as the R3clone spoligotype family (R3-family)

| **ID** | **Country** | **spoligotype** |
| --- | --- | --- |
| R3(1) | Rwanda | 1001111111111111111111111111111100001110111 |
| R3(2) | Rwanda | 1011111010011111111111111111111100001110111 |
| R3(3) | Rwanda | 1011111111111111111111111111111100001110111 |
| R3(4) | Rwanda | 0000000000000000000000001111111100001110111 |
| R3(5) | Rwanda | 1011111111011111111111111111111100001110111 |
| R3(6) | Rwanda | 1101110111111111111111111111111100001110111 |
| R3(7) | Rwanda | 1111110111011111111111111111111100001110111 |
| R3(8) | Rwanda | 1111110111111111111111111111111100001110111 |
| R3(9) | Rwanda | 1111111100001111111111111111111100001110111 |
| R3(10) | Rwanda | 1111111110011111111111111111111100001110111 |
| R3(11) | Rwanda | 1111111110111111111111111111111100001110111 |
| R3(12) | Rwanda | 1111111111011111111111111111111100001110111 |
| R3(13) | Rwanda | 1111111111110111111111111111111100001110111 |
| R3(14) | Rwanda | 1111111111111101111111111111111100001110111 |
| R3(15) | Rwanda | 1111111111111110111111111111111100001110111 |
| R3(16) | Rwanda | 1111111111111111011111111111111100001110111 |
| R3(17) | Rwanda | 1111111111111111111101111111111100001110111 |
| R3(18) | Rwanda | 1111111111111111111111111111101100001110111 |
| R3(19) | Rwanda | 1111111111111111111111111111110100001110111 |
| R3(20) | Rwanda | 1111111111111111111111111111111100001100000 |
| R3(21) | Rwanda | 1111111111111111111111111111111100001110011 |
| R3(22) | Rwanda | 1111111111111111111111111111111100001110111 |
| R3(23) | Rwanda | 1111101111111111111111111111111100001110111 |

Table S2. Overview of R3clone-specific SNPs. The Rwanda dataset was surveyed for SNPs which segregate R3clone from all others. SNPs are listed with their genomic position, the nucleotide difference between groups, and both the H37Rv name and the general gene name, where known.

| SNP position | WT (non-R3clone) | R3clone | Gene Rv name | Gene name |
| --- | --- | --- | --- | --- |
| 25631 | C | G | Rv0020c-Rv0021c (intergenic) | - |
| 98628 | G | T | Rv0090 | - |
| 102095 | C | T | Rv0092 | *ctpA* |
| 204455 | C | T | Rv0173 | *lprK* |
| 539246 | G | T | Rv0450c | *mmpL4* |
| 640643 | C | T | Rv0550c | *vapB3* |

SNPs: single-nucleotide polymorphisms. R3clone: Rwanda rifampicin-resistant clone.

Figure S1. Phylogenetic tree of the 375 isolates belonging to the R3clone cluster and their country of origin.

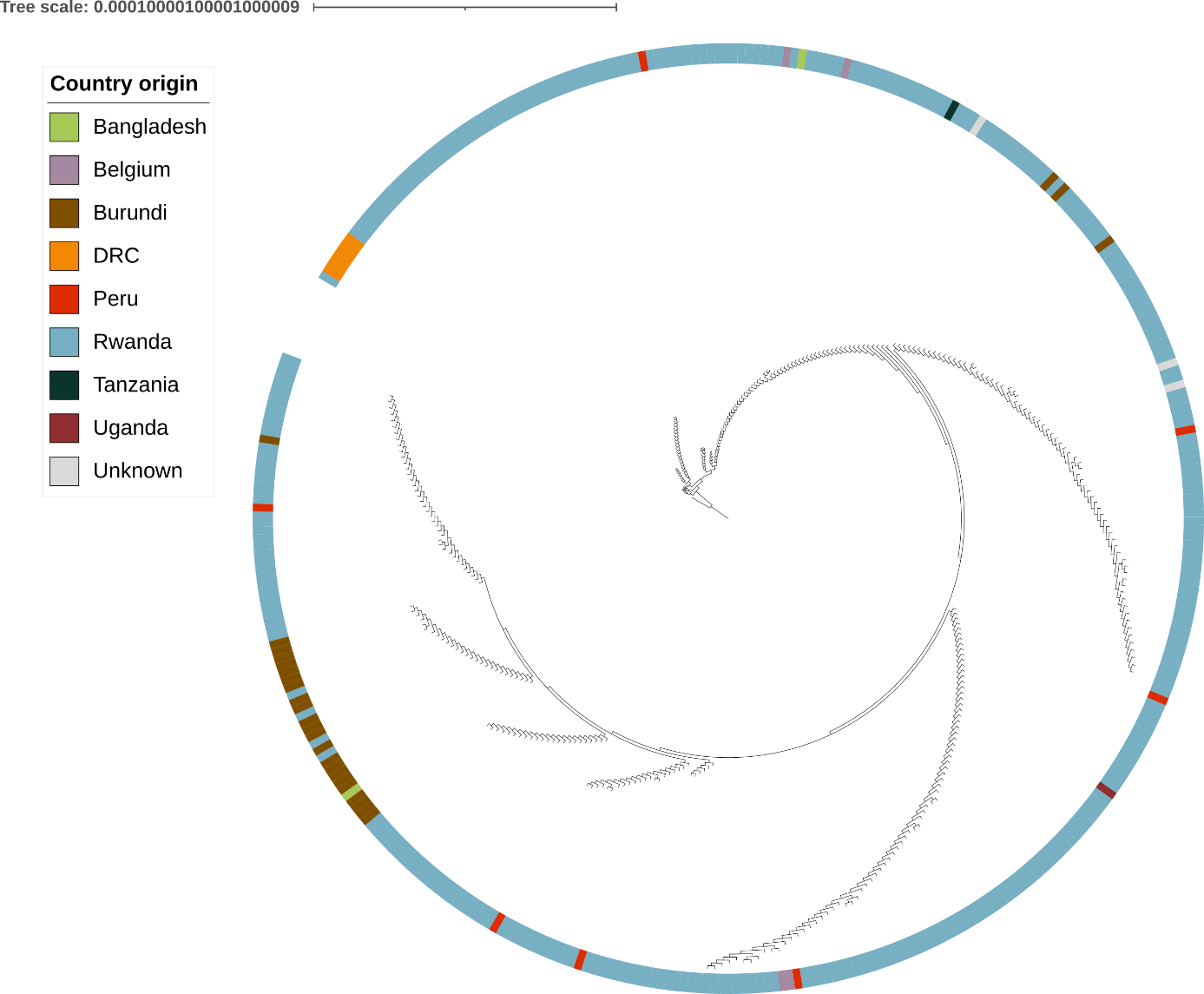
